# Association between sensitivity to the five basic tastes and frailty and low muscle mass and/or strength

**DOI:** 10.64898/2025.12.02.25341508

**Authors:** Minoru Kouzuki

## Abstract

**Aim:** Taste decline may contribute to malnutrition, frailty, and reduced muscle health in older adults, but evidence is limited and inconsistent. This study aimed to examine the association between frailty and low muscle mass and/or strength and taste-related information obtained through assessing sensitivity to the five basic tastes and using questionnaire surveys.

**Methods:** Sixty-eight community-dwelling older individuals without diagnosed taste disorders were enrolled. Assessments included background factors, sensitivity to the five basic tastes using taste-impregnated filter papers, body measurements, physical function, and a questionnaire (subjective taste, palatability, and dietary balance). Because taste sensitivity evaluation was newly devised, two scoring methods were used: (1) one point per correct answer and (2) one point for correctly identifying either low or high concentration strip for each taste.

**Results:** When using scoring method 2, it was found that a higher umami sensitivity score was associated with a significantly lower likelihood of pre-frailty or frailty (adjusted odds ratio = 0.26, p = 0.040). However, no correlation was found between muscle mass and/or muscle strength and the results of the taste sensitivity assessment. Participants reporting a decline in taste function had significantly lower umami sensitivity scores in scoring methods 1 and 2 (p=0.042, p=0.032, respectively).

**Conclusions:** Although the results slightly varied depending on the scoring method used for the taste sensitivity assessment, higher umami sensitivity may be associated with a lower risk of developing pre-frailty or frailty. Umami sensitivity was also correlated with subjective taste assessment. Changes in taste perception may affect dietary intake and cause frailty; however, longitudinal studies are warranted to establish causal relationships.

## Introduction

Frailty, sarcopenia, and dynapenia are increasingly recognized as major contributors to adverse health outcomes in older adults.^1–12^ Poor nutritional status is a contributing factor to frailty and decreased muscle mass and strength.^13^ Older individuals are prone to loss of appetite and malnutrition. The underlying causes of anorexia due to aging may be related to changes in the secretion and peripheral action of appetite hormones, effects on gastrointestinal motility, chewing and swallowing difficulties, and loss of acuity in taste.^14^ In other words, a decline in taste function, which contributes to malnutrition, can theoretically be considered a risk factor for frailty and decreased muscle mass and strength, but this association has not been fully verified. For example, a systematic review conducted to examine the association between frailty and sarcopenia and sensory disorders noted that there was insufficient information available on the association with taste disorders because of limited studies.^15,16^ In studies that assessed taste function using questionnaires and examined its association with frailty, a correlation was reported,^17,18^ but there are concerns that taste function may not have been accurately assessed. In contrast, two studies that conducted verification showed no association between frailty or sarcopenia and taste function assessed objectively using taste tests.^18,19^ However, these studies only used specific taste solutions. Sweet, salty, sour, bitter, and umami are the five basic tastes, but previous studies have not evaluated overall taste perception, suggesting that additional comprehensive evaluation of overall taste perception is warranted.

Therefore, the aim of this study was to examine the association between taste-related information obtained through an assessment of sensitivity to the five basic tastes and a questionnaire survey and frailty and low muscle mass and/or strength.

## Methods

### Participants

Participants were recruited from clubs and community gatherings in Kotoura Town (Tottori Prefecture, Japan) between September and October 2024. The inclusion criteria were aged ≥65 years at the time of consent. The exclusion criteria were diagnosis of a taste disorder with a clear cause and allergic reactions to seasonings and food additives used in the taste sensitivity assessment. Finally, 68 individuals were enrolled.

This study was approved by the Ethics Committee of the Tottori University Faculty of Medicine (No. 24A047). Prior to conducting this study, the participants were informed of the study’s aims, and written consent was obtained.

### Measurement items

#### Participant demographic characteristics

Age, sex, years of education, number of drugs taken, and medical history (hypertension, dyslipidemia, and diabetes mellitus) were investigated using a self-administered questionnaire.

#### Assessment of gustatory function

Briefly, to determine the effect of gustatory function on eating habits, individuals were evaluated on their ability to perceive tastes at concentrations detectable by most individuals in daily life. Based on the methods of previous studies,^20^ for the test, salt-coated test paper SALSAVE® (AdvanTech Toyo Co., Ltd., Tokyo, Japan) was used. The tips of the blank test strips (containing no salt) were coated with a taste solution prepared by dissolving the sample in distilled water and then dried. The concentrations of the taste solutions are listed in Table 1. Based on prior studies,^20–22^ two concentrations were prepared for each taste: a high concentration detectable by >90% of individuals without taste disorders and a lower concentration below that density. The low-concentration taste solutions were the same concentration used in previous studies,^20–22^ which was one level lower than the high-concentration solutions. Based on the results of previous studies,^20,21^ sweetness, saltiness, sourness, and umami are tastes that can be distinguished by approximately 70–80% or more of individuals. However, a previous study set the concentration for bitterness to a level that was perceived by approximately 50%,^22^ resulting in a lower correct answer rate than for other tastes. Although the other tastes were generally diluted twofold, bitterness had a lower concentration of 0.003 g/mL than with a high concentration of 0.02 g/mL. As no other studies were available for reference, I also decided to apply a twofold dilution standard to bitterness to increase the correct answer rate.

**Table 1.** Taste solution details.

| Taste | Taste substance (sales company) | Concentration (g/mL) |  |
| --- | --- | --- | --- |
| Sweet | Granulated sugar (Mitsui DM Sugar Co., Ltd.) | Low | 0.1 |
|  |  | High | 0.2 |
| Salt | Refined salt (Kyushu Salt Corporation) | Low | 0.04 |
|  |  | High | 0.1 |
| Umami | Monosodium L-glutamate (Marugo Corporation) | Low | 0.1 |
|  |  | High | 0.25 |
| Sour | Citric acid (Marugo Corporation) | Low | 0.09 |
|  |  | High | 0.165 |
| Bitter | Caffeine (Kanto Chemical Co., Inc.) | Low | 0.01 |
|  |  | High | 0.02 |

All samples used were commercially available seasonings and food additives that are safe for consumption.

The participants were instructed to place the tip of the test strip on their tongue and taste it for 3 s before removing it from their mouth. They were subsequently instructed to choose from one of seven answers: sweet, salty, sour, bitter, umami, unidentifiable taste, and no taste. The test strips were administered in the following sequence: sweet, salt, umami, sour, and bitter, and for each taste, a low-concentration was tested followed by a high-concentration. In addition, to remove any lingering taste, the participants were instructed to rinse their mouths with water before licking another test strip.

Regarding the assessment of the test, considering participants who could recognize the taste at low concentrations but not at high concentrations, the following two patterns were used to score the test results: (1) for each taste, recognition at a low concentration earns one point, and recognition at a high concentration also earns one point (each test strip counts as one point, with a maximum score of 10 points); (2) if a taste could be recognized at either low or high concentration, it was given a score of 1 (1 taste was given 1 point, and the score was out of 5 points).

#### Body measurements

Height was measured using a tape measure attached to the wall, and weight and muscle mass were measured using a weight and body composition monitor with dual-frequency bioelectrical impedance analysis (RD-800 or RD-801; TANITA Corporation, Tokyo, Japan).^23^ From the results, the body mass index (BMI) (BMI=weight [kg]/ height^2^ [m^2^]) and appendicular skeletal muscle mass index (SMI) (SMI=arm and leg skeletal muscle mass [kg]/height^2^ [m^2^]) were computed.

#### Assessment of physical functions

Physical function tests included evaluation of grip strength and gait speed. The grip strength was evaluated using a hand dynamometer (GRIP-D; T.K.K.5401; SANKA Co., Ltd., Niigata, Japan). Measurements were conducted once on each side in the standing position, and the results from the strongest hand were used in the present analysis. A three-axis accelerometer (AYUMI EYE; Waseda Elderly Health Association Co., Ltd., Tokyo, Japan) was used to measure gait speed. Gait speed was assessed by measuring the walking time at 6 m and calculating the gait speed for 1 s. To reproduce a situation close to normal walking, the participants were instructed to walk 8 m with an additional 1 m before and after the 6-m measurement point.

#### Assessment of frailty

Physical frailty was assessed using the revised Japanese version of the Cardiovascular Health Study criteria.^24^ These criteria comprised five domains: shrinking, low activity, exhaustion, weakness, and slowness. Weakness was evaluated using hand grip strength, whereas slowness was judged based on gait speed, and the remaining components were assessed via questionnaires. Participants with none of these components were considered robust, those with one or two components were considered pre-frailty, and those with three or more components were considered frailty.^23^

#### Assessment of sarcopenia and dynapenia

Sarcopenia and dynapenia were diagnosed based on the Asian Working Group for Sarcopenia (AWGS) 2019 criteria and Yamada et al.^23,25,26^ Muscle functional decline was defined as a grip strength of <28 kg in men or <18 kg in women and/or gait speed of <1.0 m/s, and reduced muscle mass was defined as an SMI of <7.0 kg/m^2^ in men or <5.7 kg/m^2^ in women. Sarcopenia was defined as low muscle mass and low muscle function, pre-sarcopenia was defined as low muscle mass without low muscle function, dynapenia was defined as low muscle function without low muscle mass, and normal was defined as anything other than the above.

### Questionnaire survey

A questionnaire survey was conducted on subjective taste function, degree of perceived palatability, and dietary balance.

To assess subjective taste function, the participants were asked “Do you feel like your sense of taste is declining?” and were instructed to choose from four options: “I feel it very much,” “I feel it a little,” “I do not feel it much,” and “I do not feel it at all.” To assess the degree of perceived palatability, the participants were asked “Are you enjoying your meals?” and were instructed to choose from four options: “I do not always find them delicious,” “I often do not find them delicious,” “I usually find them delicious,” and “I find them delicious every time.” To assess dietary balance, the participants were asked “Are you eating a balanced diet (a variety of foods)?” and were instructed to choose from four options: “It is never well-balanced,” “It is often not well-balanced,” “It is usually well-balanced,” and “It is well-balanced every day.”

Questionnaire results were analyzed by categorizing responses into positive and negative groups. Positive responses were defined as follows: for subjective taste function, “I do not feel it much” and “I do not feel it at all”; for the degree of perceived palatability, “I usually find them delicious” and “I find them delicious every time”; and for dietary balance, “It is usually well-balanced” and “It is well-balanced every day.” All other responses were classified as negative.

### Statistical analyses

Statistical analyses were performed using SPSS (version 27; IBM Corporation, Tokyo, Japan) and EZR (version 1.55; Saitama Medical Center, Jichi Medical University, Saitama, Japan), a graphical user interface for R (The R Foundation for Statistical Computing, Vienna, Austria).^27^ All statistical significance tests were two-sided, and an alpha level of 0.05 was statistically significant.

Univariate and multivariate binary logistic regression analyses were performed to examine the association between taste-related information (the independent variables) obtained from the assessment of sensitivity to the five basic tastes and questionnaire surveys and the assessment results of frailty and sarcopenia/dynapenia (the dependent variable). Because fewer individuals were assessed for frailty or sarcopenia, it is necessary to consider that the validity of the logistic model becomes problematic when the ratio of the number of events per variable analyzed is small.^28^ Therefore, frailty was classified as robust or not, and sarcopenia was classified as normal or not according to the AWGS 2019 criteria,^25^ that is, those with no decline in muscle mass and/or strength or those with a decline in muscle mass and/or strength. Furthermore, with reference to a previous study,^29^ for multivariate modeling, propensity score adjustment was used for age, sex, BMI, number of drugs taken, and medical history (hypertension, dyslipidemia, and diabetes mellitus). The propensity score was estimated using multivariate binary logistic regression analysis, with the results of the taste-related information as the dependent variable and the above adjustment factor as the independent variable. In logistic regression analysis, unadjusted and adjusted odds ratios (ORs) and 95% confidence intervals (CIs) were calculated. For the results of sensitivity of taste in all scoring methods 1 and in the total score of scoring method 2, the results were not binary; hence, a propensity score could not be calculated using binary logistic regression analysis. Therefore, a multivariate analysis was not performed. Furthermore, when comparing the results of the five basic taste sensitivity assessments between the two groups divided by the taste-related questionnaire results, the Shapiro–Wilk test was used to test for normality. In cases of normal distribution, Levene’s test was used to test for equality of variance, followed by Student’s t-test. In cases where the distribution was not normal, the Mann–Whitney U test was used.

## Results

### Characteristics of the participants

The participants’ data are presented in Table 2. The mean age was 78.3±5.8 years, and 88.2% were women. Regarding frailty status, 36 (52.9%) participants were pre-frailty and 9 (13.2%) participants were frailty. Regarding sarcopenia and dynapenia, 21 (33.3%) participants had dynapenia, 3 (4.8%) had pre-sarcopenia, and 6 (9.5%) had sarcopenia. Regarding the questionnaire results, 18 (26.5%) participants reported a decline in taste function, 8 (11.8%) participants did not find their meals tasty, and 17 (25.0%) participants felt that their meals were unbalanced. Regarding the taste sensitivity assessment, both scoring methods 1 and 2 showed lower mean scores for sweetness, umami, and bitterness than mean scores for saltiness and sourness.

**Table 2.** Characteristics of the participants.

|  | All participants (n=68) |
| --- | --- |
| Age (years) | 78.3±5.8 |
| Sex |  |
| Male | 8 (11.8) |
| Female | 60 (88.2) |
| Number of drugs taken | 2.6±2.3 |
| Medical history |  |
| Hypertension | 37 (54.4) |
| Dyslipidemia | 21 (30.9) |
| Diabetes mellitus | 7 (10.3) |
| Body mass index (kg/m<sup>2</sup>)<sup>a</sup> | 22.8±3.1 |
| Grip strength (kg) |  |
| Male | 32.5±7.1 |
| Female | 20.8±3.7 |
| Gait speed (m/sec) | 1.1±0.3 |
| Skeletal muscle mass index (kg/m<sup>2</sup>)<sup>b</sup> |  |
| Male | 7.5±0.9 |
| Female | 6.5±0.6 |
| Frailty status<sup>c</sup> |  |
| Robust | 23 (33.8) |
| Pre-frailty | 36 (52.9) |
| Frailty | 9 (13.2) |
| Sarcopenia and dynapenia staging<sup>b</sup> |  |
| Normal | 33 (52.4) |
| Dynapenia | 21 (33.3) |
| Pre-sarcopenia | 3 (4.8) |
| Sarcopenia | 6 (9.5) |
| Do you feel like your sense of taste is declining? |  |
| I feel it very much | 0 (0) |
| I feel it a little | 18 (26.5) |
| I do not feel it much | 29 (42.6) |
| I do not feel it at all | 21 (30.9) |
| Are you enjoying your meals?<sup>c</sup> |  |
| I do not always find them delicious | 3 (4.4) |
| I often do not find them delicious | 5 (7.4) |
| I usually find them delicious | 22 (32.4) |
| I find them delicious every time | 38 (55.9) |
| Are you eating a balanced diet (a variety of foods)? |  |
| It is never well-balanced | 2 (2.9) |
| It is often not well-balanced | 15 (22.1) |
| It is usually well-balanced | 39 (57.4) |
| It is well-balanced every day | 12 (17.6) |
| Taste sensitivity evaluation: scoring method 1 (point)<sup>d</sup> |  |
| Sweet | 0.93±0.70 |
| Salt | 1.24±0.71 |
| Umami | 0.97±0.88 |
| Sour | 1.41±0.70 |
| Bitter | 0.72±0.71 |
| Total score | 5.26±2.28 |
| Taste sensitivity evaluation: scoring method 2 (point)<sup>e</sup> |  |
| Sweet | 0.72±0.45 |
| Salt | 0.84±0.37 |
| Umami | 0.60±0.49 |
| Sour | 0.88±0.32 |
| Bitter | 0.57±0.50 |
| Total score | 3.62±1.30 |

Data are presented as means±standard deviations or numbers (%).
<sup>a</sup>Sample size is 67.
<sup>b</sup>Sample size is 63 (six male/57 female).
<sup>c</sup>The total does not equal 100% because of rounding to the second decimal place.
<sup>d</sup>If a taste can be recognized at both low and high concentrations, each concentration is evaluated as one point.
<sup>e</sup>If a taste can be recognized at either low or high concentration, it is evaluated as one point.

### Association between frailty and low muscle mass and/or strength with sensitivity assessment to the five basic tastes and results of taste-related questionnaires

Table 3 shows the results of the logistic regression analysis investigating the association between frailty and low muscle mass and/or strength using various assessment results. No significant association was observed between frailty and taste sensitivity assessment using scoring method 1. However, using scoring method 2, a higher umami sensitivity score was significantly associated with lower development of pre-frailty or frailty (adjusted OR=0.26, 95% CI=0.07–0.94, p=0.040). However, no statistically significant association was observed between low muscle mass and/or strength and taste sensitivity, regardless of the scoring method. Regarding the taste-related questionnaire, no significant association was observed between frailty or low muscle mass and/or strength and any of the questionnaire items.

**Table 3.**
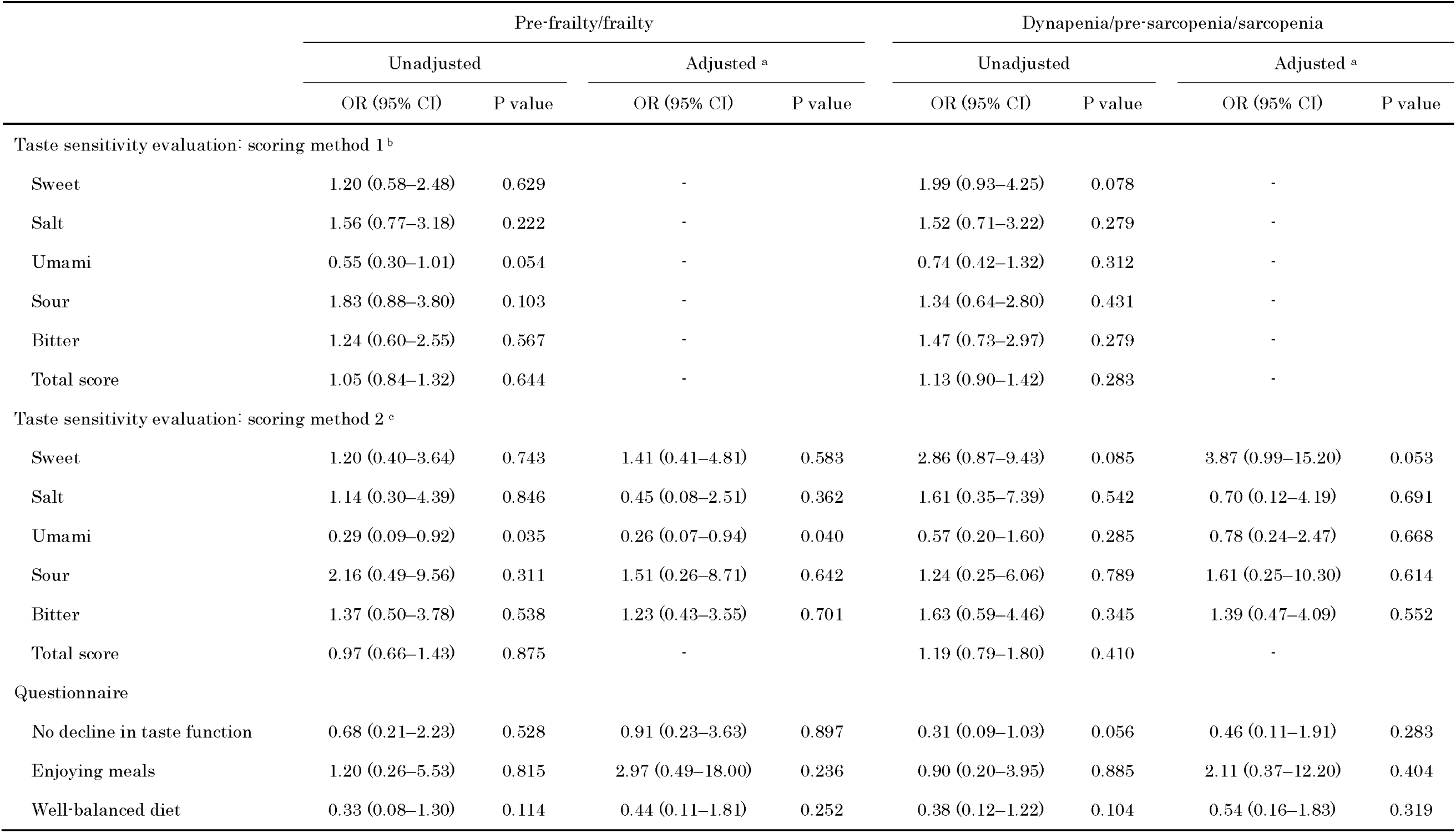
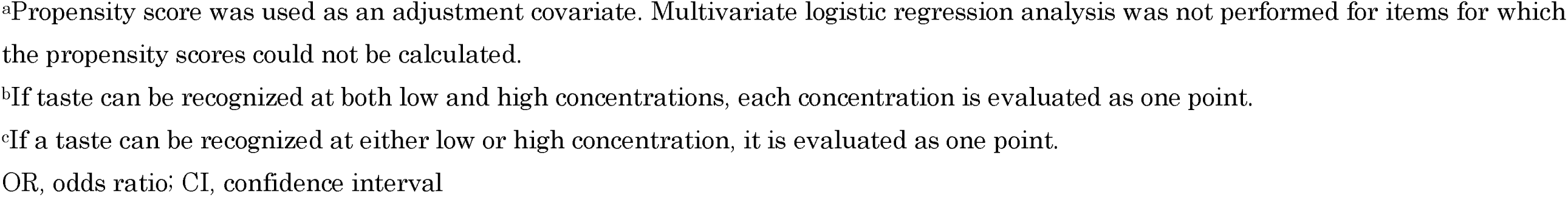
Association between the evaluation of sensitivity to the five basic tastes and information on taste obtained through questionnaire surveys and the evaluation results of pre-frailty/frailty and dynapenia/pre-sarcopenia/sarcopenia.

|  | Pre-frailty/frailty |  |  |  | Dynapenia/pre-sarcopenia/sarcopenia |  |  |  |
| --- | --- | --- | --- | --- | --- | --- | --- | --- |
|  | Unadjusted |  | Adjusted <sup>a</sup> |  | Unadjusted |  | Adjusted <sup>a</sup> |  |
|  | OR (95% CI) | P value | OR (95% CI) | P value | OR (95% CI) | P value | OR (95% CI) | P value |
| Taste sensitivity evaluation: scoring method 1 <sup>b</sup> |  |  |  |  |  |  |  |  |
| Sweet | 1.20 (0.58–2.48) | 0.629 | · |  | 1.99 (0.93–4.25) | 0.078 | · |  |
| Salt | 1.56 (0.77–3.18) | 0.222 | · |  | 1.52 (0.71–3.22) | 0.279 | · |  |
| Umami | 0.55 (0.30–1.01) | 0.054 | · |  | 0.74 (0.42–1.32) | 0.312 | · |  |
| Sour | 1.83 (0.88–3.80) | 0.103 | · |  | 1.34 (0.64–2.80) | 0.431 | · |  |
| Bitter | 1.24 (0.60–2.55) | 0.567 | · |  | 1.47 (0.73–2.97) | 0.279 | · |  |
| Total score | 1.05 (0.84–1.32) | 0.644 | · |  | 1.13 (0.90–1.42) | 0.283 | · |  |
| Taste sensitivity evaluation: scoring method 2 <sup>c</sup> |  |  |  |  |  |  |  |  |
| Sweet | 1.20 (0.40–3.64) | 0.743 | 1.41 (0.41–4.81) | 0.583 | 2.86 (0.87–9.43) | 0.085 | 3.87 (0.99–15.20) | 0.053 |
| Salt | 1.14 (0.30–4.39) | 0.846 | 0.45 (0.08–2.51) | 0.362 | 1.61 (0.35–7.39) | 0.542 | 0.70 (0.12–4.19) | 0.691 |
| Umami | 0.29 (0.09–0.92) | 0.035 | 0.26 (0.07–0.94) | 0.040 | 0.57 (0.20–1.60) | 0.285 | 0.78 (0.24–2.47) | 0.668 |
| Sour | 2.16 (0.49–9.56) | 0.311 | 1.51 (0.26–8.71) | 0.642 | 1.24 (0.25–6.06) | 0.789 | 1.61 (0.25–10.30) | 0.614 |
| Bitter | 1.37 (0.50–3.78) | 0.538 | 1.23 (0.43–3.55) | 0.701 | 1.63 (0.59–4.46) | 0.345 | 1.39 (0.47–4.09) | 0.552 |
| Total score | 0.97 (0.66–1.43) | 0.875 | · |  | 1.19 (0.79–1.80) | 0.410 | · |  |
| Questionnaire |  |  |  |  |  |  |  |  |
| No decline in taste function | 0.68 (0.21–2.23) | 0.528 | 0.91 (0.23–3.63) | 0.897 | 0.31 (0.09–1.03) | 0.056 | 0.46 (0.11–1.91) | 0.283 |
| Enjoying meals | 1.20 (0.26–5.53) | 0.815 | 2.97 (0.49–18.00) | 0.236 | 0.90 (0.20–3.95) | 0.885 | 2.11 (0.37–12.20) | 0.404 |
| Well-balanced diet | 0.33 (0.08–1.30) | 0.114 | 0.44 (0.11–1.81) | 0.252 | 0.38 (0.12–1.22) | 0.104 | 0.54 (0.16–1.83) | 0.319 |

<sup>a</sup>Propensity score was used as an adjustment covariate. Multivariate logistic regression analysis was not performed for items for which the propensity scores could not be calculated.
<sup>b</sup>If taste can be recognized at both low and high concentrations, each concentration is evaluated as one point.
<sup>c</sup>If a taste can be recognized at either low or high concentration, it is evaluated as one point.
OR, odds ratio; CI, confidence interval

### Association between sensitivity assessment for the five basic tastes and taste-related questionnaire results

In scoring methods 1 and 2 of the taste sensitivity assessment, the umami sensitivity scores were significantly lower in participants who reported a decline in taste function (p=0.042 and p=0.032, respectively) (Table 4). However, no significant association was found between the taste sensitivity assessment and questionnaire results regarding the degree of perceived palatability or dietary balance.

**Table 4.** Comparison of taste-related questionnaire results and sensitivity assessment of the five basic tastes.

|  | Do you feel like your sense of taste is declining? |  |  | Are you enjoying your meals? |  |  | Are you eating a balanced diet (a variety of foods)? |  |  |
| --- | --- | --- | --- | --- | --- | --- | --- | --- | --- |
|  | Yes | No | P value | No | Yes | P value | No | Yes | P value |
| Taste sensitivity evaluation: scoring method 1 (point) <sup>a</sup> |  |  |  |  |  |  |  |  |  |
| Sweet | 1.11±0.58 | 0.86±0.73 | 0.175 | 1.00±0.53 | 0.92±0.72 | 0.715 | 1.06±0.66 | 0.88±0.71 | 0.356 |
| Salt | 1.11±0.83 | 1.28±0.67 | 0.478 | 1.12±0.64 | 1.25±0.73 | 0.570 | 1.00±0.71 | 1.31±0.71 | 0.110 |
| Umami | 0.61±0.85 | 1.10±0.86 | 0.042 | 0.88±0.83 | 0.98±0.89 | 0.752 | 0.94±0.75 | 0.98±0.93 | 0.898 |
| Sour | 1.22±0.81 | 1.48±0.65 | 0.239 | 1.12±0.83 | 1.45±0.67 | 0.252 | 1.35±0.79 | 1.43±0.67 | 0.801 |
| Bitter | 0.72±0.75 | 0.72±0.70 | 0.970 | 0.75±0.89 | 0.72±0.69 | 0.983 | 0.76±0.75 | 0.71±0.70 | 0.793 |
| Total score | 4.78±2.53 | 5.44±2.19 | 0.111 | 4.88±2.85 | 5.32±2.22 | 0.611 | 5.12±2.15 | 5.31±2.35 | 0.762 |
| Taste sensitivity evaluation: scoring method 2 (point) <sup>b</sup> |  |  |  |  |  |  |  |  |  |
| Sweet | 0.89±0.32 | 0.66±0.48 | 0.065 | 0.88±0.35 | 0.70±0.46 | 0.304 | 0.82±0.39 | 0.69±0.47 | 0.278 |
| Salt | 0.72±0.46 | 0.88±0.33 | 0.122 | 0.88±0.35 | 0.83±0.38 | 0.765 | 0.76±0.44 | 0.86±0.35 | 0.345 |
| Umami | 0.39±0.50 | 0.68±0.47 | 0.032 | 0.62±0.52 | 0.60±0.49 | 0.893 | 0.71±0.47 | 0.57±0.50 | 0.320 |
| Sour | 0.78±0.43 | 0.92±0.27 | 0.111 | 0.75±0.46 | 0.90±0.30 | 0.220 | 0.82±0.39 | 0.90±0.30 | 0.388 |
| Bitter | 0.56±0.51 | 0.58±0.50 | 0.858 | 0.50±0.53 | 0.58±0.50 | 0.657 | 0.59±0.51 | 0.57±0.50 | 0.888 |
| Total score | 3.33±1.24 | 3.72±1.33 | 0.141 | 3.62±1.60 | 3.62±1.28 | 0.767 | 3.71±1.21 | 3.59±1.34 | 0.888 |

Data presented as means±standard deviations.
<sup>a</sup>If a taste can be recognized at both low and high concentrations, each concentration is evaluated as one point.
<sup>b</sup>If a taste can be recognized at either low or high concentration, it is evaluated as one point.

## Discussion

Although the association between frailty and general taste sensitivity was not found, there may be a possible association between high umami sensitivity and lower development of pre-frailty or frailty. This study represents the first evaluation of the association between umami sensitivity and frailty status has not yet been investigated in previous studies.^18,19^

In this study, the mechanisms linking frailty to decreased taste sensitivity could not be investigated. However, according to previous studies, a possible mechanism is that a decline in taste function affects dietary intake, leading to frailty. First, aging affects taste function. Although some studies have documented an association between aging and decreased taste function, there is no consensus regarding which taste sensitivity declines with age.^30,31^ However, taste sensitivity does not improve with age, and older individuals may experience a decline in taste sensitivity due to some factors.^32,33^ This suggests that older individuals are at risk for changes in taste sensitivity due to several factors. Regarding the impact of changes in taste on dietary intake, a previous study has shown that individuals with subjective symptoms of taste disorders have a higher frequency of appetite disorders than those without.^34^ Furthermore, loss of appetite contributes to frailty.^35,36^ The presence or absence of loss of appetite were not evaluated; thus, this is the only speculation. However, because participants with subjectively impaired taste function had significantly lower umami sensitivity than those without, it is possible that decreased taste sensitivity led to perceived changes in the taste of food, which in turn affected dietary intake. However, no correlation was found between taste sensitivity and simple questions such as whether the food was tasty or well-balanced. Therefore, further investigations are required to determine how changes in taste sensitivity affect dietary intake. Moreover, as this was a cross-sectional study, further studies are required to establish causal associations.

Lower muscle mass and/or strength was not associated with taste sensitivity, consistent with the result of a previous study.^19^ However, similar to the mechanism underlying frailty, it is possible that changes in taste function affect dietary intake, leading to decreased muscle mass and strength. Certainly, previous studies have reported associations between appetite or the ability to eat and body composition and muscle strength.^37^ In this regard, for example, high dietary diversity is associated with lower cognitive and physical frailties, low sleep disturbance and mental disorders, and good nutritional status.^38^ Moreover, healthy dietary patterns were associated with higher quality of life.^39^ When considering the impact of changes in dietary intake due to decreased taste sensitivity, it may be best to consider it not only as a physical factor (decreased muscle mass and strength) but also as a factor affecting a wide range of mental, psychological, and nutritional aspects, potentially increasing the risk of health problems. In other words, although decreased taste sensitivity is a factor in health problems, its impact on individual health components may be limited.

Finally, this study has some limitations. First, the sample size was small (n=68). When the participants were divided into groups based on the assessment of frailty and sarcopenia/dynapenia, the number of participants in each group was small, making it impossible to adjust for background factors in some analyses. Furthermore, the assessment of background factors, which are related to frailty, sarcopenia, and dynapenia,^40–42^ was insufficient. Second, since the sense of taste could not be accurately assessed, the results should be interpreted with caution. This study entailed an evaluation of whether perceived tastes of most individuals could be identified. Whether individuals can perceive faint concentrations of taste or whether they experience pathological taste disorders has not yet been evaluated.

In conclusion, although the results slightly varied depending on the scoring method used to assess taste sensitivity, these findings suggest that a higher umami sensitivity may be associated with a lower prevalence of pre-frailty or frailty. Furthermore, because umami sensitivity is also associated with subjective taste function, it is possible that changes in food taste may affect food intake and lead to frailty, although further verification is required to prove this causal association. Given that 26.5% of the participants reported a decline in taste function and 25.0% reported an unbalanced diet, it is important to address the issue of preventing anorexia and malnutrition in older individuals. In a previous study, in which the authors compared the perceived intensity and pleasantness of four series of food items, consisting of five geometrically-spaced concentration levels, it was found that older adults had higher optimal preferred perceived intensities than did younger individuals.^43^ One of the reasons that older individuals prefer stronger flavors is a change in taste function. For example, some studies have shown that adding seasonings or sauces increases food intake.^44–46^ Adjusting seasoning can be considered a method to enhance palatability and appetite. However, caution should be exercised regarding excessive salt intake. Nutritional education focused on the use of spices and herbs in a diet has been shown to have the potential of improving several indicators of diet quality and attitudes toward healthy eating.^47^ Therefore, providing information on how to adjust seasonings may also improve dietary imbalances. In futures studies, it is anticipated that further efforts focusing on taste perception will be undertaken for the prevention of frailty.

## Author contributions

MK designed the study, implemented the tests, analyzed and interpreted the data, wrote the first draft of the manuscript, and reviewed the manuscript.

## Acknowledgements

I would like to thank all the individuals who participated in this study and express my gratitude to the staff of the Health Division in Kotoura Town and all others who cooperated with the study. I also thank Editage (www.editage.com) for the English language editing and their assistance in creating the graphical abstract for this paper.

## Funding statement

This study was supported by the 30th Umami Research Grant from the Society for Research on Umami Taste. The funding body had no role in study design, data collection and analysis, decision to publish, or preparation of the manuscript.

## Conflict of interest disclosure

MK has received a research grant and travel expenses from the Society for Research on Umami Taste.

## Data availability statement

The datasets used and/or analyzed in the current study are available upon reasonable request from the corresponding author, with the approval of the ethics committee.

## Ethics approval and patient consent statements

The ethical review board of the Faculty of Medicine, Tottori University (Yonago, Japan) approved the design of this study (Number: 24A047), which was conducted in accordance with the tenets of the Declaration of Helsinki. Prior to conducting this study, the participants were informed of the study’s aims, and written consent was obtained.

## Notes

### Summary of Updates

Updated the "Funding statement" and "Conflict of interest disclosure"

